# Forecasting undetected COVID-19 cases in Small Island Developing States using Bayesian approach

**DOI:** 10.1101/2020.05.13.20100545

**Authors:** Andrio Adwibowo

## Abstract

In dealing with the COVID-19, the fundamental question is how many actually undetected cases are going around regarding the capabilities of current health systems to contain the virus?. Due to a large number of asymptomatic cases, most COVID-19 cases are possibly undetected. For that reason, this study aims to provide an efficient, versatile, easy to compute, and robust estimator for the number of undetected cases using Bayes theorem based on the actual COVID-19 cases. This theorem is applied to 25 Small Island Developing States (SIDS) due to SIDS vulnerability. The results in this study forecast that possibly undetected COVID-19 cases are approximately 4 times larger than the numbers of actual COVID-19 cases as observed. This finding highlights the importance of using modeling tool to get the better and comprehensive of current COVID-19 cases and to take immediately precaution approaches to mitigate the growing numbers of COVID-19 cases as well.

## 1. Introduction

Recently, mathematical models have been widely used to forecast COVID-19 based on the probability of moving between states from susceptible to infected states and then to a recovered states or death for example (Chen *et al*. 2020, Menkir *et al*. 2020). Mathematical model (Dawson *et al*. 2015, Aronis *et al*. 2020) is a common versatile tool to identify what it needs to be done in order to deal the challenge. Models are useful to estimate the extent of the epidemic reduction in the effective transmission rate required to control an epidemic in a specific manner. There are several mathematical models have been used to forecast COVID-19 that include Poisson distribution and Bayesian approach. As an example, a study by Bayes *et al*. (2020) has used a Bayesian approach and a prior epidemic COVID-19 history to forecast the total number of COVID-19 related deaths in Peru for the next seventy days. The advantage of Bayesian approach because it can solve an uncertainty and analyze whether a mitigation measure is being effective and forecast the worst-case scenarios. Bayesian approach incorporates predictive posterior distribution in epidemiological models aims to analyze a full spectrum of scenarios and enable to determine whether the response is adequate in order to prevent the downfall of the health system.

Nowadays, COVID-19 cases per May 2020 have reached million cases spreading in more than 100 countries worldwide. Most of the cases are observed in the landlocked countries, mainland and continents. Nonetheless, the case analyses in Small Island Developing States (SIDS) are still limited. According to UN-OHRLLS, SIDS are a distinct group of developing countries facing specific social, economic and environmental vulnerabilities. Those SIDS are spread over 3 main geographical regions namely the Caribbean, the Asia Pacific, and the Africa. UN lists there are approximately 50 SIDS in those regions. The SIDS are facing common challenges including narrow resource, small domestic markets, heavy dependence on a few external and remote markets, high costs for energy, infrastructure, transportation, communication, little resilience to natural disasters, growing populations, and fragile natural environments. Therefore, SIDS are highly disadvantaged in the development process and require special support from the international community.

Another emerging challenge faced by SIDS is health and communicable disease issues. SIDS countries with their small, geographically disparate populations combined with limited health workforces are particularly vulnerable to the burden of diseases (Kranenburg and Essink, 2015). In the current COVID-19 pandemic, like states in the mainland and continents, SIDS are also experiencing this pandemic. The COVID-19 source databases provided by Johns Hopkins Corona Resource Center and Worldometer may have reportedly the cases in SIDS in comprehensive manner. Nonetheless, there are probability of undetected COVID-19 cases as concerned by numerous studies (Mukhopadhyay and Chakraborty, 2020). Likewise in dealing with this uncertainty in COVID-19 reporting, several studies have emphasized the modeling approach, like study by Gayawan *et al*. (2020) using truncated Poisson distribution to predict COVID 19 cases in Africa countries. Nonetheless, their study focused only on the African countries located in the mainland and the data about the SIDS in Africa regions are still limited.

Considering aforementioned situation, there is an urgent need to study the current COVID 19 cases mainly in vulnerable SIDS. For that reason, this study aims to forecast the possibility of undetected COVID 19 cases in SIDS.

## 2. Methodology

### 2.1. Data collection

Data up to May 10, 2020 used in this study were collected from data repository provided by Johns Hopkins Corona Resource Center at Johns Hopkins University Center for Systems Science and Engineering (CSSE) and the Worldometer websites (Max Roser and Ortiz-Ospina, 2020). The Worldometer itself is well known free data repository and the reference website which is trusted by the UK Government and Johns Hopkins CSSE. Through these resources, numbers of COVID 19 cases from SIDS were obtained. In total there are 25 SIDS from Africa (SIDS), Caribbean (SIDS), and Asia Pacific (SIDS) have been investigated in this study. The numbers of sampled SIDS from Africa, Caribbean, and Asia Pacific are 4, 18, and 3 respectively.

### 2.2. Bayesian approach

An analysis to estimate the undetected COVID-19 cases is Bayes theorem following Bayes *et al*. (2020) and Yamagata (2020). This theorem is as follows:

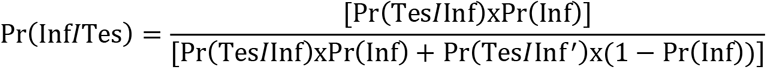

The following probability events and notations are considered:

Pr(Inf): is the prior probability (%) of COVID-19 cases in a population.

Pr(Tes/Inf): is the probability (%) of a positive COVID-19 test result given for positive COVID-19. Pr(Tes/Inf’): is the probability (%) of a positive COVID-19 test result given for negative COVID-19.

Pr(Inf/Tes): is the probability (%) of an undetected COVID-19 case.

Assuming that all SIDS follow the aforementioned model relation between the Covid-19 tests and confirmed cases, a rough estimate of the number of undetected COVID-19 cases Pr(Inf/Tes) in each SIDS can be made.

## 3. Results

Figure 1 shows numbers of Small Island Developing States (SIDS) that have 0–50, 51–100, and 101–400 actual COVID-19 cases. High SIDS were observed for 0–50 COVID-19 cases classes or accounted for up to 30% of total countries with 0–50 COVID-19 cases. The highest SIDS percentage was observed under 51–100 classes even though the SIDS numbers were low. The smallest percentage and number of SIDS were observed when the COVID-19 cases were increased or under 101–400 COVID-19 cases classes.

**Figure 1.**
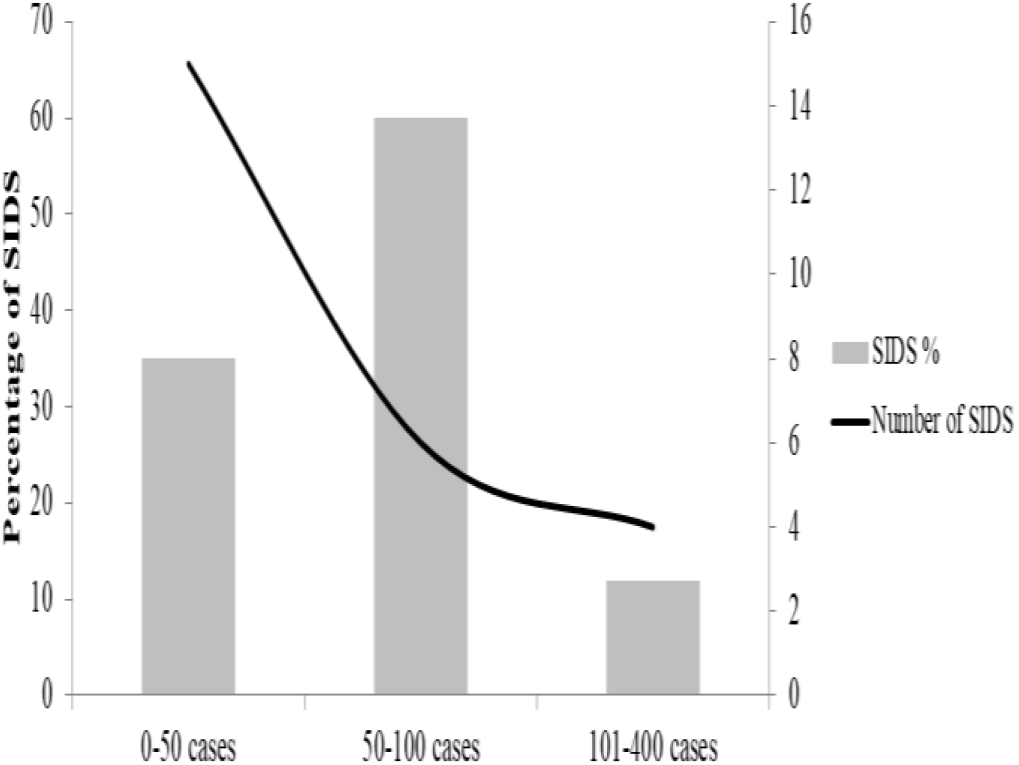
The percentage and number of SIDS for 0–50, 51–100, and 101–400 actual COVID-19 cases classes.

Figure 2 shows numbers of SIDS that have 0–50, 51–100, and 101–400 actual COVID-19 cases classified into SIDS regions. Numbers of SIDS in Caribbean were always high for each cases class. SIDS in Asia Pacific were having lower actual COVID-19 cases compared to SIDS in Caribbean and Africa. While African SIDS have higher actual COVID-19 cases in comparison to SIDS in Asia Pacific and Caribbean. In average the SIDS in Asia Pacific has the lowest actual COVID-19 cases. As can be seen in Table 1, the mean of actual COVID-19 cases for Asia Pacific, Caribbean, and Africa are 32 (95%CI:20.8–43.2), 43.7 (95%CI: 42.6- 44.8), and 140.5 (95%CI: 14.2–295.2) respectively.

**Figure 2.**
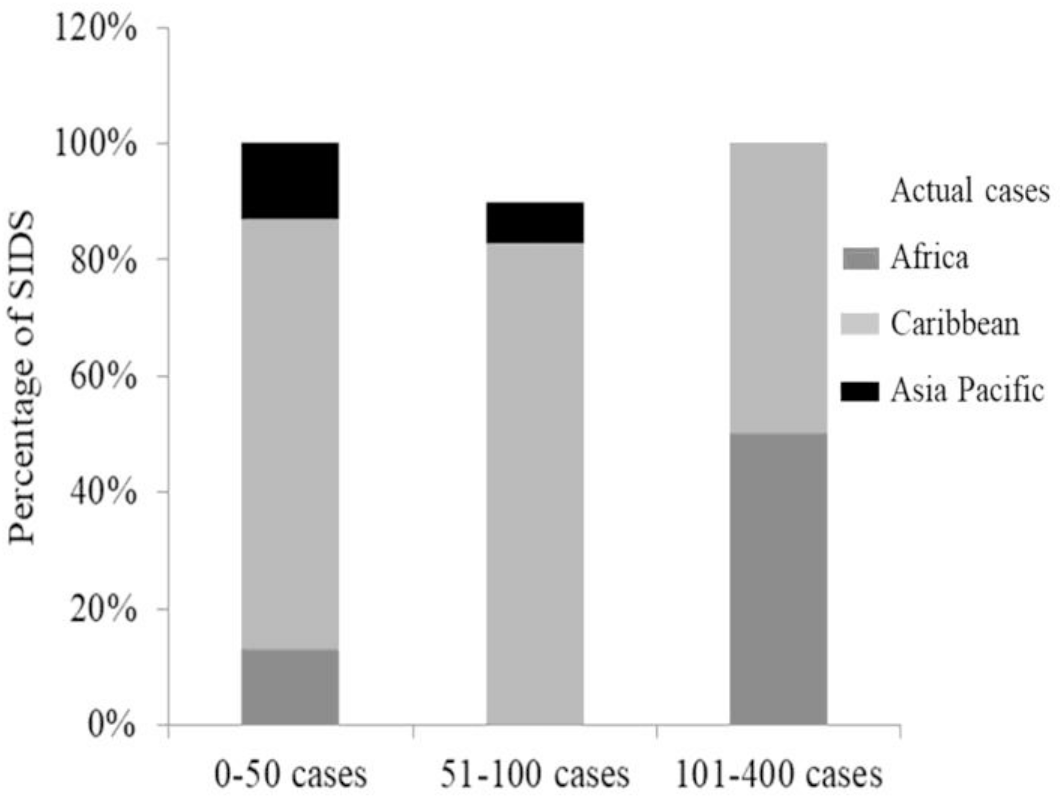
The percentage of SIDS for 0–50, 51–100, and 101–400 actual COVID-19 cases classes according to SIDS regions.

**Table 1.** Actual and undetected Covid-19 cases for several SIDS regions at May 10, 2020.

| Cases | Regions | Mean | SD | Lower 95%CI | Upper 95%CI |
| --- | --- | --- | --- | --- | --- |
| Actual | Africa | 140.5 | 157.8 | 14.2 | 295.2 |
|  | Caribbean | 43.7 | 2.3 | 42.6 | 44.8 |
|  | Asia Pacific | 32 | 24.2 | 20.8 | 43.2 |
| Possibly undetected | Africa | 512.2 | 613.7 | 89.2 | 1113.6 |
|  | Caribbean | 144.2 | 3.6 | 142.5 | 145.9 |
|  | Asia Pacific | 137.2 | 96.4 | 92.6 | 181.7 |

The following plots in Figure 3 and 4 shows the correlation of population with both the total actual and possibly undetected COVID-19 cases in SIDS regions. This suggests a clear estimation that the undetected cases for SIDS in Africa and Caribbean compared to Asia Pacific are about 2 to 4 times as() large (Figure 4) as compared to the total actual cases. The mean of possibly undetected COVID-19 cases for Asia Pacific, Caribbean, and Africa are 137.2 (95%CI: 92.6–181.7), 144.2 (95%CI: 142.5- 145.9), and 512.2 (95%CI: 89.2–1113.6) correspondingly (Table 1).

**Figure 3.**
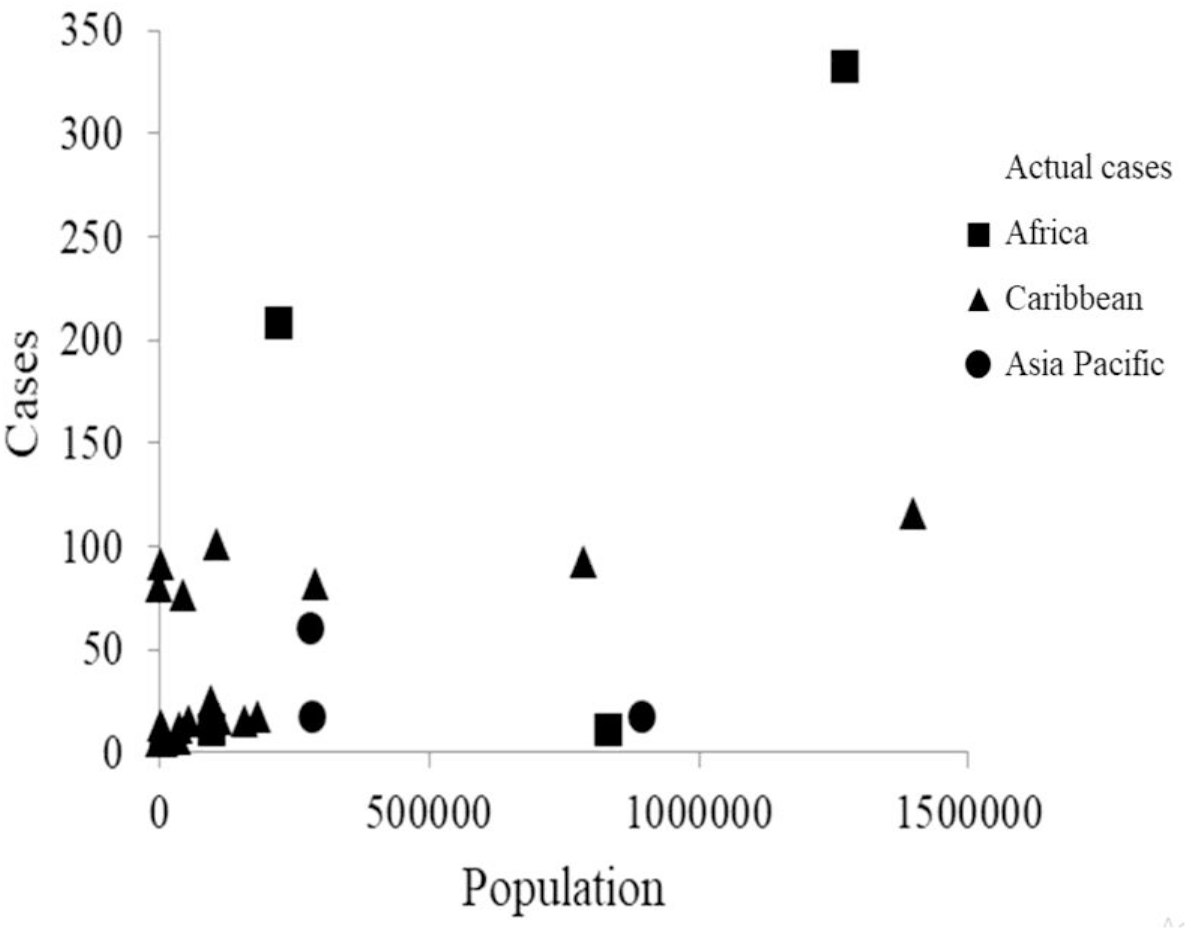
The correlation plots between population with actual COVID-19 cases of SIDS in Africa, Caribbean, and Asia Pacific regions.

**Figure 4.**
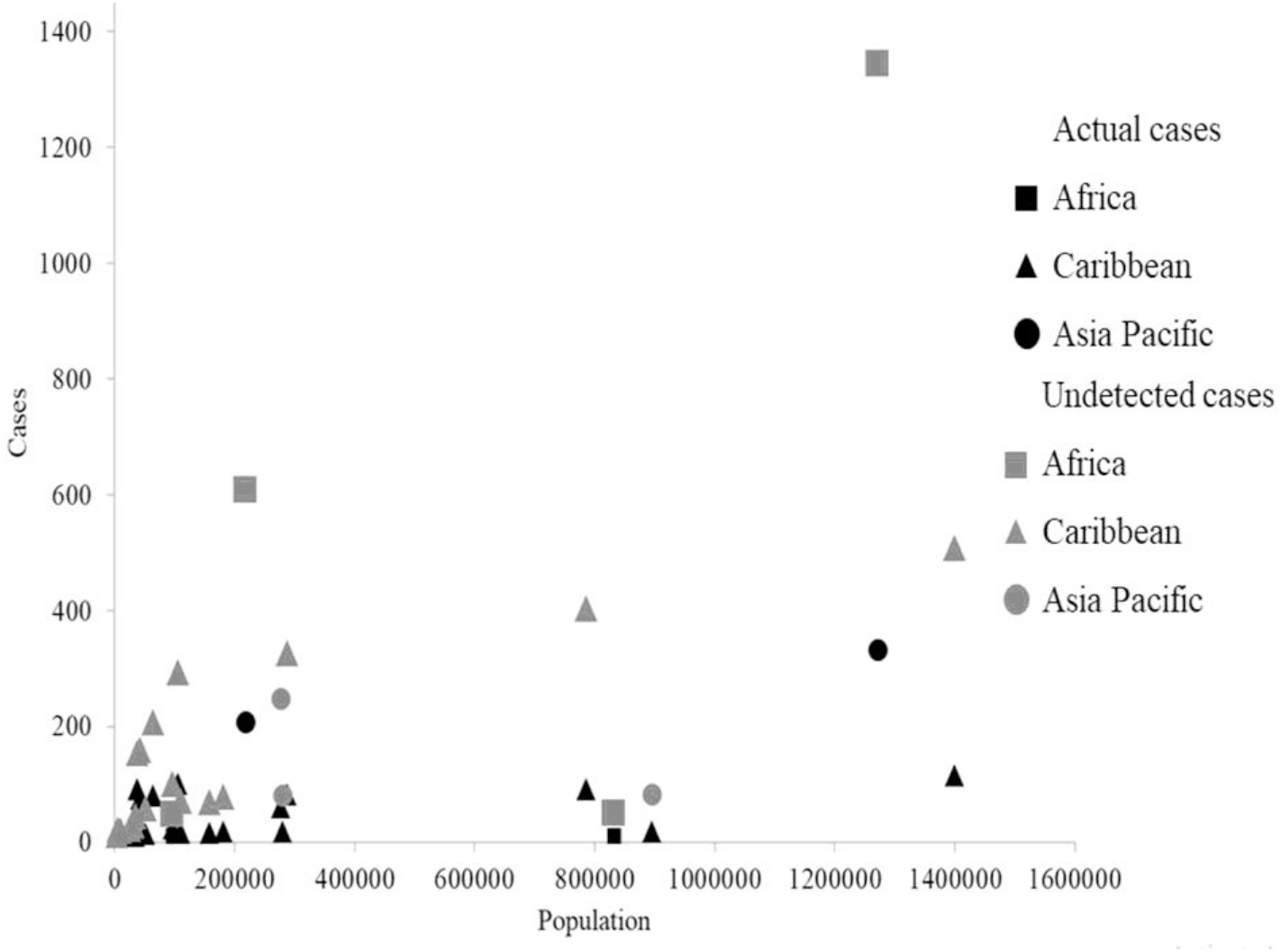
The correlation plots between population with actual COVID-19 and possibly undetected cases of SIDS in Africa, Caribbean, and Asia Pacific.

## 4. Discussions

The use of Bayesian as predictive tools to forecast COVID 19 is growing rapidly as can be seen in the number of literatures published recently. The forecasted results provided show whether there are increasing or declining trends from the actual data. In his research, Yamagata (2020) applied a Bayesian approach to estimate daily changes of reproduction number (R_o_) to determine the infection speed and reporting rate as an effect of health policy. The result showed declining trend of R_o_ indicating the COVID-19 cases were affected by policy. Nonetheless in some situations, the data are poor or incomplete (Mena *et al*. 2020). Bayesian approach is considered as a way to circumvent the data incompleteness. This approach is also a versatile tool to incorporate observations and use the full range of parameter estimates contained in the posterior distribution to adjust for uncertainties in model predictions.

Data incompleteness led to undetected COVID-19 cases are emerging challenges in dealing with this epidemic (Stock *et al*. 2020). One way to deal with this situation is by applying quantification modeling. By using model, Pedersen and Meneghini (2020) predict that in Italy, at the beginning of the epidemic the number of unidentified SARS-nCov2-positive individuals was equal to 10 times the number of confirmed cases. In this study, it is forecasted that undetected COVID-19 cases in SIDS is around 4 times the number of actual cases

The main interesting findings from this study are despite the actual COVID-19 cases per May 2020 have reached up to a million cases around the globe, nonetheless there are several countries that still have actual COVID-19 cases less than 100 cases. This study recorded that numerous SIDS are those countries that still having low COVID-19 cases during 4 months of pandemic durations. Most particular SIDS in Pacific regions are still having low actual COVID-19 cases as well. One of the major concerns is the systematic uncertainty in the number of population who have hosted the virus. The major contribution to this uncertainty is possibly due to the small fraction of Covid-19 tests being performed.

Correspondingly, the proposed Bayesian approach offered in this study aims to answer the essential question which is how many actually undetected cases are going around among SIDS regarding the capabilities of SIDS health systems to contain the virus?. Of course, this study has succeeded to estimate the possible numbers of undetected cases, nonetheless those numbers are advised to be used as a starting point each time an intervention and approach to deal with the COVID-19 pandemic are delivered. Bayes theorem as used in this study is versatile and easy to apply in practice and this is one of the merits of the approach. The another fundamental advantage of using Bayes theorem to forecast the undetected COVID-19 is lies on the data feed. In this study, the required data are simply use time series of confirmed cases data which is readily and freely available from government endorsed trusted open sources.

In fact the numbers of COVID-19 related deaths per 100000 populations in SIDS are higher than numbers observed in least developed countries, landlocked developing countries, Africa, and Southeast Asia regions. The presence of COVID-19 in SIDS is raising a concern regarding the SIDS vulnerability and fragility. Based on the past experiences, a communicable disease has threatened SIDS. In 2019, there were several SIDS have experienced a measles outbreak which killed 81 and infected more than 5600 people. This epidemic forced those SIDS into a six-week-long state of emergency. Currently, there are several SIDS in Caribbean region for instance are experiencing COVID-19 case fatality rates as high as many hardest hit countries in Europe regions. The high prevalence of pre-existing comorbidities including diabetes, cardiovascular diseases, and obesity, make populations in SIDS particularly vulnerable to COVID-19 (Nicole *et al*. 2018). In fact, available literature provides evidence suggests that populations having with these comorbidities are more likely to develop severe symptoms from COVID-19. Corresponding to the SIDS vulnerability, most SIDS are having economy situations that are lacking capacities for detection and treatment of a novel disease. Likewise, those SIDS are lacking health capabilities and ability to treat the sick and protect health workers.

In this study, SIDS in Africa regions are having notably a fourfold increase of COVID-19 undetected cases compared to Caribbean and Asia Pacific regions. This related to the COVID-19 exportation cases from the China. As reported by Menkir *et al*. (2020), several countries in Africa and including SIDS have received the most imported cases from China ranging from 0.4 to 3.0 cases respectively. The numbers of undetected COVID-19 cases presented in here should provide early warning. As the comparison between the actual cases (Figure 3) and undetected estimations (Figure 4) shows that more tests are required to capture the possibly undetected cases, thus now is the high time to raise the testing efficiency in order to reduce the undetected COVID-19 cases. This seems to be the only good way to reduce the death rate of COVID-19 patients as indicated by the large amount of COVID-19 testing in Germany and South Korea for examples. Likewise, this can warn the SIDS to take immediately precaution approaches to mitigate the growing numbers of COVID-19. Those precaution steps including prevention, detection, and rapid responses to epidemic issues.

## 5. Conclusions

The finding presented in this study just a first evidence on the use of Bayesian approach to forecast COVID-19 data in particular SIDS. With Bayesian approach, this study forecasted that the numbers of undetected cases are approximately 4 times larger than the numbers of actual cases. Bayesian theorem used in this study provides a straightforward solution to shed light on undetected cases by incorporating heterogeneity that may arise in the probability of being detected.

## 6. Recommendations

The finding from this study regarding the estimation of the undetected Covid-19 cases can provide recommendations for authorities to plan economic policies, make decisions around different stages of lockdown if necessary, and to work towards the production of intensive health care systems. Likewise, there is still a lot of room for improvement. As for recommendations and future works, there are a number of avenues that can be explored including theoretically from the modeling perspective and as a predictive tool. For instance, incorporating the SIR model to Bayesian approach would be an option required for improvement. For reliable results, improvement of robustness would be required as well.

## Data Availability

Data used in this study were collected from data repository.

https://www.worldometers.info/coronavirus

## Notes

### Competing Interest Statement

The authors have declared no competing interest.

